# Epileptogenicity alters intrahippocampal ripple propagation

**DOI:** 10.64898/2026.06.13.26355594

**Authors:** Yihe Chen, Honyi Ye, Lingqi Ye, Lingli Hu, Cong Chen, Richard Staba, Shuang Wang, Shennan Aibel Weiss

## Abstract

**Objective:** Tracing the propagation of high-frequency oscillations (HFOs) aids in localizing epileptogenic regions and improving surgical outcomes. We examined how hippocampal epileptogenicity influences the propagation properties of the HFOs it generates.

**Methods:** We analyzed non-REM sleep stereo-EEG from 49 patients (68 hemispheres) with verified hippocampal contacts. Hippocampi were stratified by excitability: 28 seizure onset zone (SOZ), 22 more-irritative non-SOZ (>6 interictal epileptiform discharges [IED]/min), and 18 less-irritative non-SOZ (<6 IED/min). To isolate significant HFO propagation pathways, we constructed empirical temporal networks (maximum latency 150 ms) and validated them against 1,000 permutation-generated surrogates. We then compared the proportion of statistically significant propagating HFOs originating from hippocampal contacts across these groups.

**Results:** We examined ripples on oscillation (RonO, 80-250 Hz) and fast ripples on oscillation (FRonO, 250-600 Hz). FRonO, but not RonO, rates were significantly elevated in hippocampal SOZ versus non-SOZ contacts (p<1e-9). Intrahippocampal RonO propagation proportion was highest in less-irritative non-SOZ compared to more-irritative non-SOZ (p<0.05) and SOZ (p<0.001). Across groups, we found no other differences in RonO or FRonO propagation proportions, including spread to other mesial-temporal structures or the neocortex.

**Significance:** Intrahippocampal RonO propagation is proportionally greater in less-irritative non-SOZ tissue than in the epileptogenic hippocampus. Distinguishing physiological from pathological RonO using signal features alone remains challenging. Our work suggests these categories are not distinct; rather, RonO signals and their underlying hippocampal substrates likely exist on a continuous physiological-to-pathological spectrum. Furthermore, RonO propagation may serve as a novel metric to quantify hippocampal epileptogenicity.

**Key Points:**

1. Intrahippocampal propagation of ripples on oscillations (RonO, 80–250 Hz) is highest in the less-irritative hippocampus along the epileptogenicity spectrum despite their rates were similar.
2. The proportions of hippocampal-to-mesial-temporal and hippocampal-to-neocortical RonO propagation remain constant across varying degrees of hippocampal epileptogenicity.
3. Propagation proportions for fast RonO (fRonO, 250–600 Hz) do not differ significantly across the hippocampal epileptogenicity spectrum despite that their rates differ.

**Plain Language Summary:** The hippocampus is a part of the brain that is crucial for memory. Sometimes, injured parts of the hippocampus create fast electrical brainwaves, called high-frequency oscillations (HFOs), which are linked to seizures. However, healthy parts of the hippocampus also create HFOs, and these are important for learning and memory. In this study, we wanted to know: do the brainwaves from an injured hippocampus spread differently through the brain than the brainwaves from a healthy one?

## Introduction

High-frequency oscillations (HFOs) consist of ripples (80-250 Hz) and fast ripples (250-600 Hz)^1–3^. When a ripple or a fast ripple is generated superimposed on the EEG background (i.e., a slower oscillation) we refer to them as RonO or FRonO, respectively. In animal models of epilepsy^4–11^ and in patients with drug resistant epilepsy^6,12–19^, undergoing pre-surgical evaluation with stereo EEG (SEEG), increased rates of RonO and FRonO detected in the LFP and intracranial EEG (iEEG) are promising biomarkers of epileptogenicity. This is particularly the case when the ripple and fast ripple are superimposed on epileptiform spikes^20,21^. However, RonO is generated in healthy brain too and play an important role in memory and cognition^22^. An open question is whether normal physiological RonO can be distinguished from pathological RonO^23–26^.

Murine studies show that hippocampal RonO are essential in spatial memory consolidation. A certain physiological RonO called a hippocampal sharp-wave ripples (SpW-R, 80-250 Hz) recorded in the local field potential (LFP), most often from area CA1, during immobility and non-REM sleep coincides with compressed place cell neuronal action potential sequence in a forward and reverse replay^2,22^. Using electrical brain stimulation to disrupt these hippocampal SpW-R during non-REM sleep decreases performance in repeat spatial navigation tasks^27^. Evidence shows that SpW-R propagates in the hippocampus at a velocity of 0.35 m/s^22,28–31^. While intrahippocampal SpW-R propagation is thought to be critical for spatial memory consolidation^32^ direct causal evidence has yet to be established in rats or mice.

Rather than assuming distinct categories of RonO without a ground truth, we investigated whether the propagation properties of hippocampal RonOs could serve as a marker to gauge their relative position on the spectrum between physiological SpW-Rs and pathological events. We defined the extent of epileptogenicity in the hippocampus as high seizure onset zone (SOZ), medium (i.e., non-SOZ and high IED rate, more irritative), and low (non-SOZ and low IED rate, less irritative), and hypothesized the severity of epileptogenicity corresponds with extent of functional and morphological disturbances that affects the propensity of RonO and FRonO to propagate.

We tested this hypothesis using intracranial EEG (iEEG) recordings, during non-REM sleep, from patients undergoing stereo-EEG (SEEG) evaluation who had recording contacts in the hippocampus. We then identified the timing of RonO and FRonO events and constructed temporal networks of their paths of propagation. Identifying the electrode contact considered as the source node of HFO propagation has been found to correspond to the epileptogenic zone that is necessary and sufficient for seizure generation^33–36^. We therefore anticipated that the proportion of RonO and FRonO that propagate would be greatest in the hippocampal SOZ.

## Materials and Methods

### 1. Data collection

We retrospectively enrolled patients with drug-resistant epilepsy who underwent SEEG in The Second Affiliated Hospital of Zhejiang University from 2019 to 2025. We included patients with following criteria: (1) Patients with unilateral or bilateral hippocampal electrodes; (2) 20-50min NREM stage2-stage3 SEEG recording at least 4 hours after seizure and 4 hours before seizure; (3) Available of pre-SEEG structural MRI scanning, post-SEEG CT scanning. The institutional review boards of Zhejiang University Hospital approved this study (IRB: IR2022189).

### 2. Identification of contacts MNI coordinates

T1-weighted structural MRI was obtained before intracranial electrode implantation and was standardized to normalized MNI space using non-linear normalization function from SPM12 implemented in the MATLAB toolbox Brainstorm (Version: 20-Dec-2023, https://neuroimage.usc.edu/brainstorm/). Post-implantation CT was then co-registered to normalized pre-implantation T1 MRI. Each contact was labeled according to hyperdense metallic signal on post-implantation CT manually and the normalized MNI coordinate of each contact was exported. Based on these coordinates, each contact was assigned to an anatomic brain region; contacts localized to white matter were excluded from further analyses.

### 3. iEEG recording and sleep recording

The implantation plan for depth electrodes (8–16 contacts, 2 mm in length, separated by 1.5 mm, and 0.8 mm in diameter; HKHS, Beijing, China) was formulated individually for each patient according to clinical hypotheses regarding seizure onset zone and propagation zone from non-invasive data. Subdermal needle electrodes were also placed at positions F3, C3, T3, P3, F4, C4, T4, P4, Fz, Cz and Pz as well as the bilateral mastoids. In each patient, SEEG signals were recorded on a 256-channel Nihon-Kohden system at a sampling rate of 2000 Hz.

To enable sleep staging, chin electromyography electrodes and electrooculography electrodes were attached to the skin 72 hours after implantation surgery. Sleep stages were scored in 30-s epochs according to the American Academy of Sleep Medicine (AASM) criteria by an experienced neurologist. For each patient, we selected a 20–50 min artifact-free segment from NREM stage N2–N3 sleep. Sleep recordings of SEEG were saved as EDF files.

### 4. HFO and interictal epileptic spike detection

For the extracted EDF data, preprocessing was performed in MATLAB toolbox eeglab (v2019.1; https://sccn.ucsd.edu/eeglab). The general preprocessing workflow was as follows: 1) Channels of interest were renamed selected using pop_chanedit and pop_select function; 2) Using the pop_eegfiltnew function, a 50-Hz notch filter was applied to remove power-line interference.

Then interictal spikes were detected from bipolar channels using MATLAB based toolbox spike_detector_hilbert_v23^37^. Ripples (80-250Hz) and fast ripples (250-600Hz) were detected automatically using a MATLAB-based Hilbert HFO detector developed in our laboratory^38^. The onset times of HFO events were identified as the earliest time point at which the amplitude reached 50% of the peak amplitude. In these analyses we excluded ripples and fast ripples that coincided with spikes. The detailed methodology of HFO detection was described in the Supplementary Materials.

### 5. Identifying and characterizing HFO propagation paths in a temporal network

We identified the intra-hemispheric HFO propagation paths from within the hippocampus, and from the hippocampus to other structures based on the time-respecting sequences of HFO events (Figure 1A-2, B1-B2). A sequence was defined if the latency between two adjacent HFO events, in two different bipolar channels, was below a specified maximum latency. We defined maximum latency as either 150msec or 32.5 msec. Some paths included multiple hops between channels, with each hop restricted by the maximum latencies. We excluded paths beginning with contacts outside the hippocampus even if they next propagated into hippocampus. Paths beginning in the hippocampus that exhibited backtracking or cycles (i.e., Contact A to Contact B then back to Contact A) were still included.

**Figure 1.**
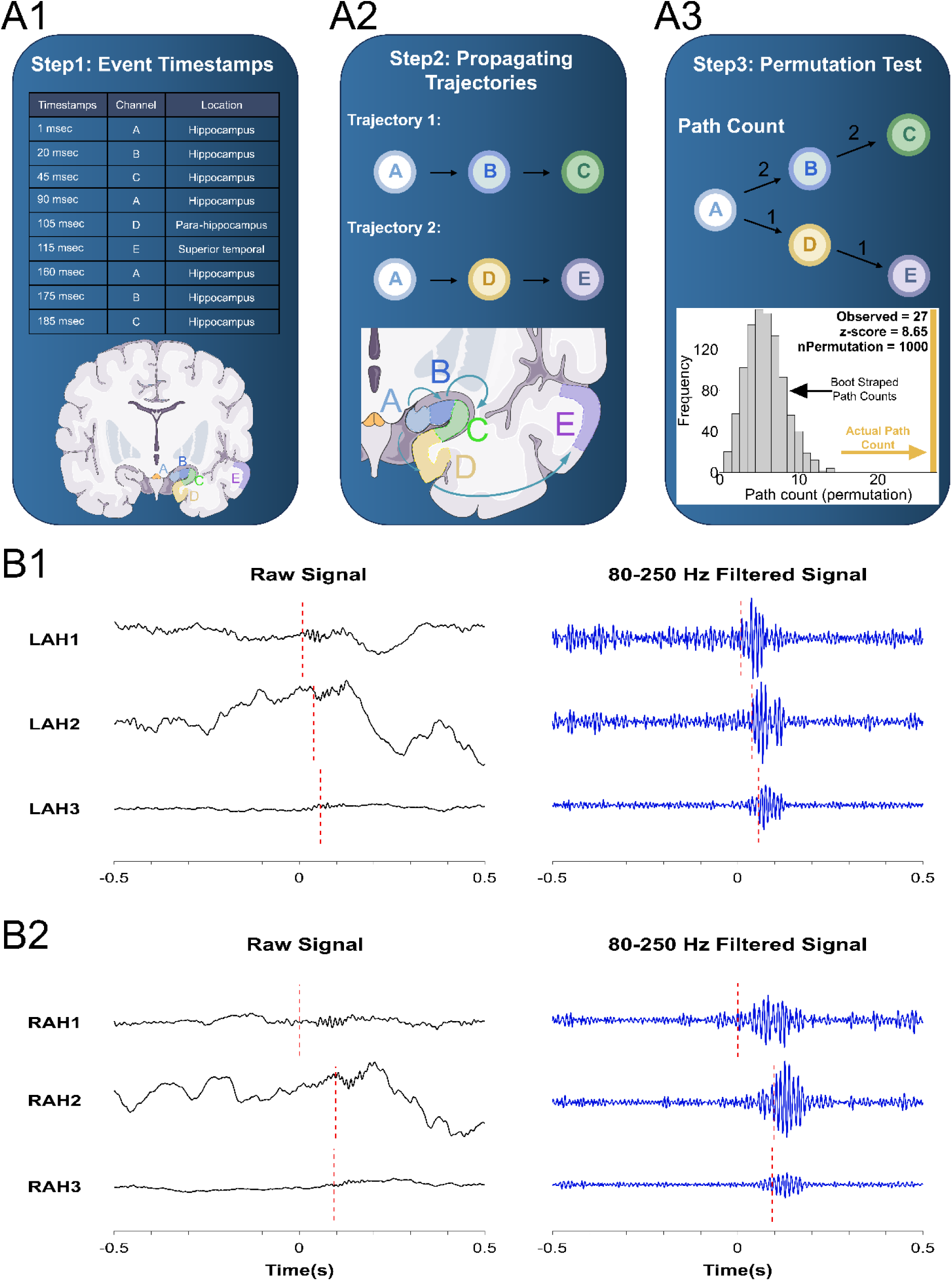
Construction and validation of HFO temporal networks and propagation paths originating from hippocampus. (A1) Step-1: Detected HFO events were organized by onset timestamps, channel label, and location (see panel below). (A2) Step-2: Events were temporally linked with a latency less than 150 msec or 32.5 msec to form time-respecting propagating trajectories across channels. The corresponding schematic brain (below) shows the spatial propagation of the HFO events. (A3) Step-3: Boot strapping permutation tests were used to compare the actual number of HFO propagation counts for a certain path with 1000 surrogates, in which the HFO onset times were randomized for each node. The actual propagation path count was compared with the propagation count of the surrogates to derive a z-score and p-value. (B1) Representative examples of RonO intra-hippocampal propagtion event with shorter latency in raw and band-pass filtered iEEG signals. Left panels show the raw signals from three hippocampal channels (LAH, left anterior hippocampus), and right panels show the corresponding band-pass filtered signals. Red dashed vertical lines indicate the onset of the RonOs. (B2) Representative examples of RonO intra-hippocampal propagtion event with longerr latency in raw and band-pass filtered iEEG signals. Left panels show the raw signals from three hippocampal channels (RAH, right anterior hippocampus), and right panels show the corresponding band-pass filtered signals. Red dashed vertical lines indicate the onset of the RonOs.

**Figure 2.**
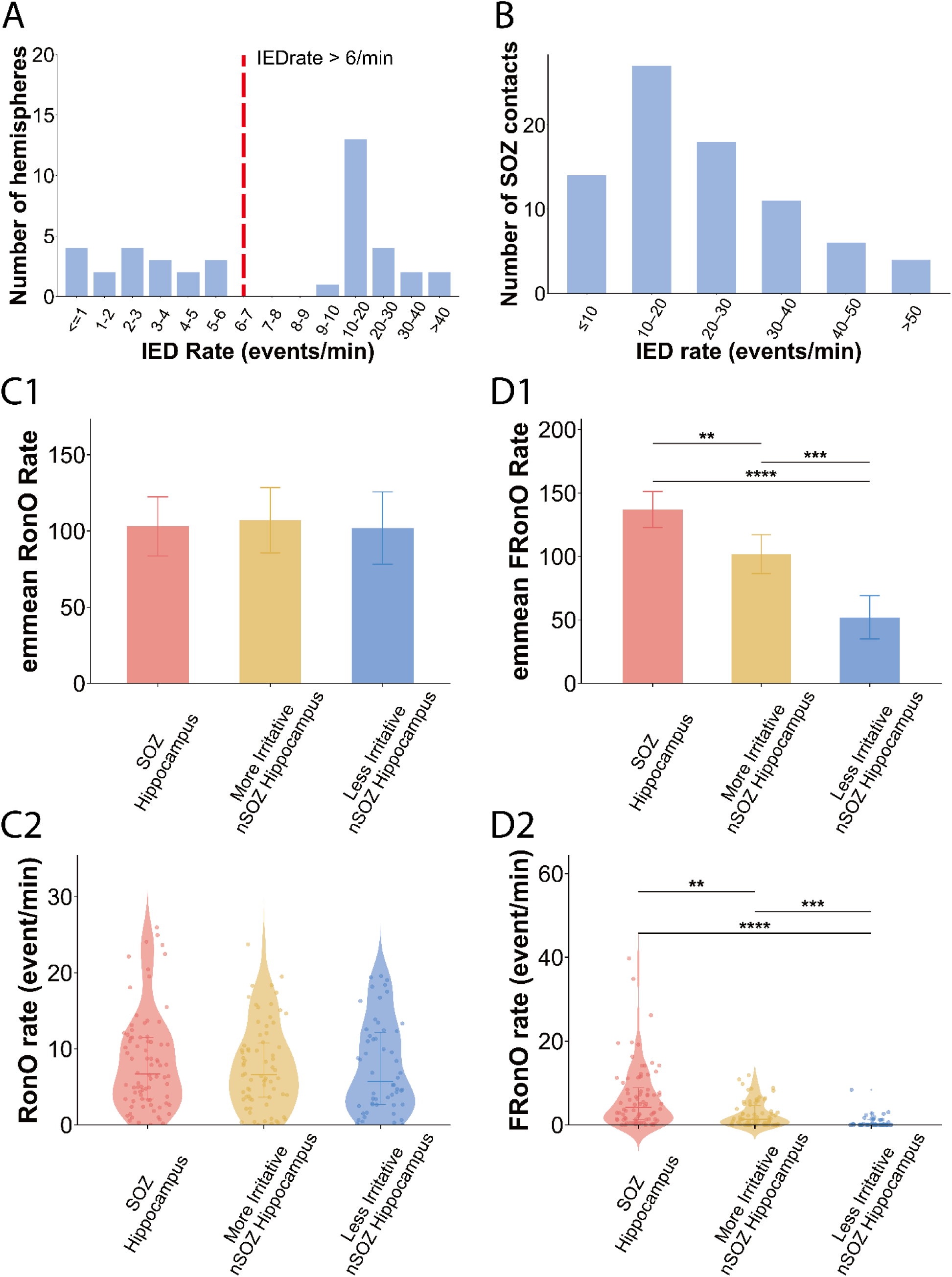
Defining the three groups of hippocampi on a spectrum of increasing epileptogenecity and comparison of RonO rate and fRonO rate across these 3 groups. (A) Within patient hemispheres, maximal hippocampal inter-ictal epileptiform discharges (IED) rates were used to distinguish less irritative hippocampi in the non-seizure onset zone (nSOZ) from more irritative hippocampi in the nSOZ. To balance sample size, a cut-off of greater of 6 IED/min (red, vertical hasehed line) was used to distinguish certain contacts in the nSOZ designated hemispheres of the more irritative hippocampus. (B) IED rates for contacts that are epileptogenic and clinically designated as the SOZ, for reference to the IED rates identified in the less and more irritative nSOZ groups in panel A, (C) RonO rate in the hippocampus across these 3 groups and (D) fRonO rate in the hippocampus across 3 groups. (C1, D1) Bars represent Aligned-rank estimated marginal mean (emmean) and error bars indicate 95% CI. (C2, D2) Violin plots of actual RonO/FRonO rates in events/min. Each dot represents one hippocampal contact. Error bars indicate the median with interquartile range, shown as the 25th to 75th percentiles. Post hoc pairwise comparisons were performed using the Holm–Bonferroni correction. Significant pairwise comparisons are shown above the bars: ***P < 0.001, ****P < 0.0001.

The time respecting paths were defined using pathpy (https://github.com/pathpy/pathpy)^39,40^ to build temporal graphs utilizing higher-order network analysis. We then asked which of these paths, defined by pathpy, were significantly more likely than chance using a 1000 surrogate-based bootstrapping framework that randomized HFO times at all the contacts (Supplementary Methods). This resulted in a decrease in false positive paths due to the relatively longer allowed maximum propagation latencies (Figure 1A3). Whereas pathpy can identify traveling wavefronts without utilizing bootstrapping, combining a longer maximum latency with boot strapping can identify poly-synaptic event cascades, and state changes.

In network science, the conditional probability of an event fully transmitting along a specific pathway is a standard metric. To ensure statistical validity, we only analyzed pathways that appeared significantly above chance levels in our bootstrapping simulations. HFO propagation proportion (or probability) for each significant path was defined over the entire duration of analyzed iEEG as:

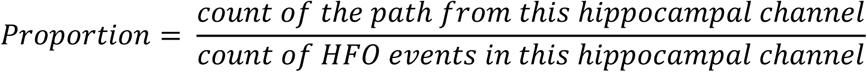

Last, to reduce dimensionality of this HFO propagation proportion analysis, the HFO propagation proportions for significant paths were grouped into 3 categories based on whether the path: 1) remained in the hippocampus (i.e., intra-hippocampal propagation); 2) propagated from the hippocampus and to other mesial-temporal lobe structures, but did not reach the neocortex (i.e., hippocampus to MTL propagation); or 3) propagated from hippocampus to a contact in the neocortex (i.e., hippocampus to neocortex propagation). Importantly, if a propagation path originating in the hippocampus reached the MTL or neocortex at any point and subsequently returned to the hippocampus, it was still classified as a hippocampus-to-MTL or hippocampus-to-neocortex path, respectively. The complexity of the temporal graph analysis was also reduced by ignoring the number of hops in the propagation paths specified by the temporal graph generated by pathpy. Temporal graphs generated by pathpy cannot easily be used to measure distance and propagation velocity. The related hop measure is highly dependent on the spatial sampling of electrode contacts in and around the hippocampus.

### 6. Patient group

Patient data was divided by hemisphere into three groups based on whether the hippocampus was designated the seizure onset zone (SOZ hippocampus, i.e., Group 1) or non-SOZ. If the hippocampus was designated non-SOZ but at least a single contact generated inter-ictal epileptiform discharges (IED) at a rate > 6/min it was labeled as more irritative non-SOZ hippocampus (i.e., Group 2), if all hippocampal contacts generated IED at a rate <6/min it was labeled as less irritative non-SOZ hippocampus (i.e., Group 3). With respect to identifying the significant paths in the temporal network for these three groups we examined only paths beginning from hippocampal contacts designated the SOZ in Group 1, contacts in the non-SOZ with an IED rate >6/min in Group 2, and all non-SOZ contacts in Group 3. Paths beginning from these certain hippocampal contacts could propagate to other hippocampal contacts unbound by this inclusion criteria.

### 7. Statistical analysis

We compared the rate (event/min) and propagation proportion of two types of hippocampal HFO: 1) ripple on background/oscillation (i.e., RonO, 80-250 Hz) and 2) fast ripple on background/oscillation (i.e., FRonO, 250-600 Hz) rates across 3 groups. The data was analyzed by aligned rank transform ANOVA using R package ARTool. Holm-Bonferroni correction was then used for post hoc pairwise comparison of aligned-rank estimated marginal means which were calculated by R package emmeans.

We used a two-way mixed-effects ANOVA to analyze the interaction effect of group and the three categories of propagation using R packages lme4 and lmerTest, followed by the Holm-Bonferroni corrected comparison of estimated marginal means to compare the proportions across 3 groups within 3 propagation categories respectively.

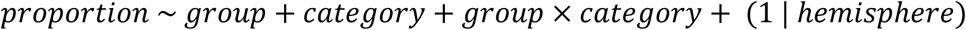

In this formula, propagation proportion was set as a response variable, while group, category and group × category were set as fixed effects while hemisphere was set as random effect. P<0.05 was considered as statistically significant. All statistical analysis were accomplished using R version 4.3.0.

## Results

### 1. Patient Demographics

We enrolled 49 patients, including 22 patients with mesial temporal lobe epilepsy with a hippocampal seizure onset zone (SOZ), 6 patients with mesial temporal epilepsy plus (with hippocampal and neocortical SOZ) and 21 patients with neocortical epilepsy (with neocortical SOZ). Of the 49 patients, 26 were male and 23 were female. The mean age was 24.7 ± 9.0 years; 33 patients were adults and 16 were pediatric (<18 years). Overall, 26 patients underwent unilateral SEEG implantation and 23 underwent bilateral implantation; among those with bilateral implantation, 19 had bilateral hippocampal electrodes.

For patients with neocortical SOZ, 11 patients had SOZ in frontal lobe, 2 patients had occipital SOZ, 3 patients had parietal SOZ, 2 patients had insular SOZ, 2 had temporal neocortical SOZ and 1 had multi-focal SOZ in temporal neocortex, insula and frontal. Surgical outcomes were assessed using the Engel classification at least 1 year after surgery. The mean follow time was 31.5 ± 16.2 months. Among the 22 patients with MTLE, 17 were seizure-free (Engel Ia–Id) and 5 were not seizure-free (Engel II–IV) after resection or ablation. Among the 6 patients with MTLE+, 5 were not seizure-free and 1 did not undergo surgery. Among the patients with neocortical epilepsy, 12 were seizure-free and 8 were not seizure-free. The mean duration of the SEEG recordings, per patient analyzed was 33.98 ± 7.96 min. On average, 11 ± 2 electrodes were implanted, and 92 ± 21 contacts were included in the analysis.

### 2. Distinguishing three groups of hippocampi on a spectrum of epileptogenicity

Group 1 hippocampi (i.e., SOZ hippocampus) were defined by the epileptologist as including at least one depth EEG contact labeled as the SOZ. To determine the cutoff for defining highly irritative non-SOZ from less irritative non-SOZ hippocampi, we first calculated the contact-level inter-ictal epileptiform discharge (IED) rate for all non-SOZ hippocampal contacts. For each hemisphere, the hippocampal contact with the highest IED rate was then identified and assigned to a predefined IED-rate bin which generated a hemisphere-level histogram of IED rates (Figure. 2A). From this histogram, a cut-off of >6 IED/min was used to balance the number of non-SOZ subject-hemispheres in Group 2 (i.e. more irritative non-SOZ hippocampus) and in Group 3 (i.e., less irritative non-SOZ hippocampus). Similar IED rates were seen in the irritative hippocampal contacts in Group 2 and the hippocampal SOZ contacts in Group 1 (Figure. 2B). In total, 68 hemispheres were included in the analysis: including 28 hemispheres in Group 1 (i.e., SOZ hippocampus), 22 hemispheres in Group 2 (i.e., more irritative non-SOZ hippocampus) and 18 in Group 3 (i.e., less irritative non-SOZ hippocampus). A comparison of the mean number of certain hippocampal contacts, such as those labeled SOZ or exhibiting >6 IED/min, used for rate and path analysis in the three groups as compared to the mean number of total hippocampal contacts is shown in Figure. 3. ANOVA demonstrated a significant difference in the number of all hippocampal contacts across the 3 groups of hippocampi (p<0.05). Post-hoc comparisons showed fewer of all contacts in the less irritative hippocampi compared to contacts in the group of hippocampi in the SOZ. However, with respect to the certain hippocampal contacts used as the source node for significant propagation paths, ANOVA demonstrated no significant difference across the 3 groups of hippocampi.

**Figure 3.**
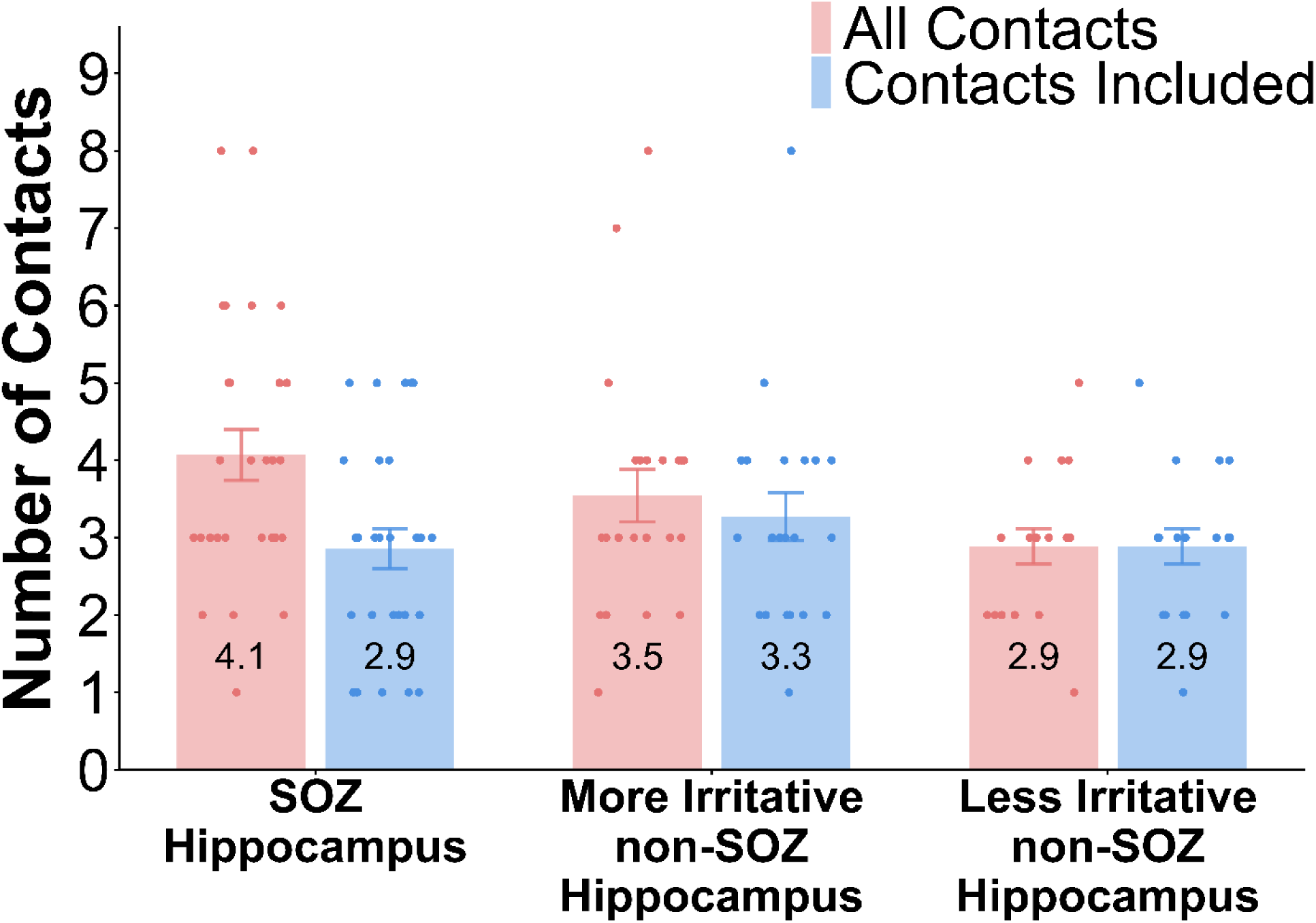
Number of total hippocampal contacts (red), and contacts included for source node of propagation paths (blue) across all hemispheres and in the three groups of hippocampi. Error bars indicate the standard error of the mean. Numbers inside the bars indicate the group mean. ANOVA demonstrated a significant difference in all contacts across the 3 groups of hippocampi (p<0.05). Post-hoc comparisons show fewer “all contacts” in the less irritative hippocampi compared to “all contacts” in the group of hippocampi in the SOZ. ANOVA demonstrated no significant difference in the number of “contacts included” across the groups of hippocampi.

### 3. Impact of Hippocampus Epileptogenicity on Hippocampal HFO Propagation

We compared RonO (i.e., ripple on oscillation/background 80-250 Hz) and fast RonO (i.e., FRonO, 250-600 Hz) rates in the hippocampi across the 3 groups. Because we used IED rate to distinguish Group 2 and 3, we did not compare the rate or propagation HFO spikes across the groups to avoid circular reasoning. Using an aligned rank transform ANOVA, we found that the RonO rates showed no significant differences across the 3 groups (p=0.95) (Figure. 2C1-2, Supplementary Table. S1). We have previously reported similar findings^41^. Then, we compared the hippocampal FRonO rate across three groups. There was significant difference of FRonO rates across 3 groups (p=1.2259e-09, Supplementary Table. S3), where SOZ hippocampi had highest FRonO rates and less irritative non-SOZ hippocampi had lowest FRonO rates in 3 groups (Figure. 2D1-2, Supplementary Table. S2).

We next compared the differences of HFO propagation proportion from the certain hippocampal contacts in the three groups using a maximum latency of 150msec with two-way mixed-effects ANOVA. We examined three categories of propagation from the hippocampus: 1) intrahippocampal propagation; 2) propagation from the hippocampus to other mesial temporal lobe structures; and 3) propagation from the hippocampus to the neocortex. There was a significant interaction effect between hippocampal group and the category of propagation (p=0.0059, Supplementary Table. S3). Pairwise comparison demonstrated that intrahippocampal propagation proportion of RonO was highest in Group 3 (i.e., less irritative non-SOZ hippocampi), as compared to Group 2 (i.e., more irritative non-SOZ hippocampi, p=0.03) and Group 1 (i.e., SOZ hippocampi, p=0.0004) (Figure. 4A). No difference was observed between the three groups with respect to propagation proportion from the hippocampus to other mesial-temporal lobe structures (Figure.4B, Supplementary Table. S3) or neocortex (Figure. 4C, Supplementary Table. S3). We also repeated this analysis and generated temporal networks with a shorter maximal latency of 32.5 msec. In this case the two-way mixed-effects ANOVA showed no significant interaction effect between group and category (Supplementary Table. S4, p=0.18).

**Figure 4.**
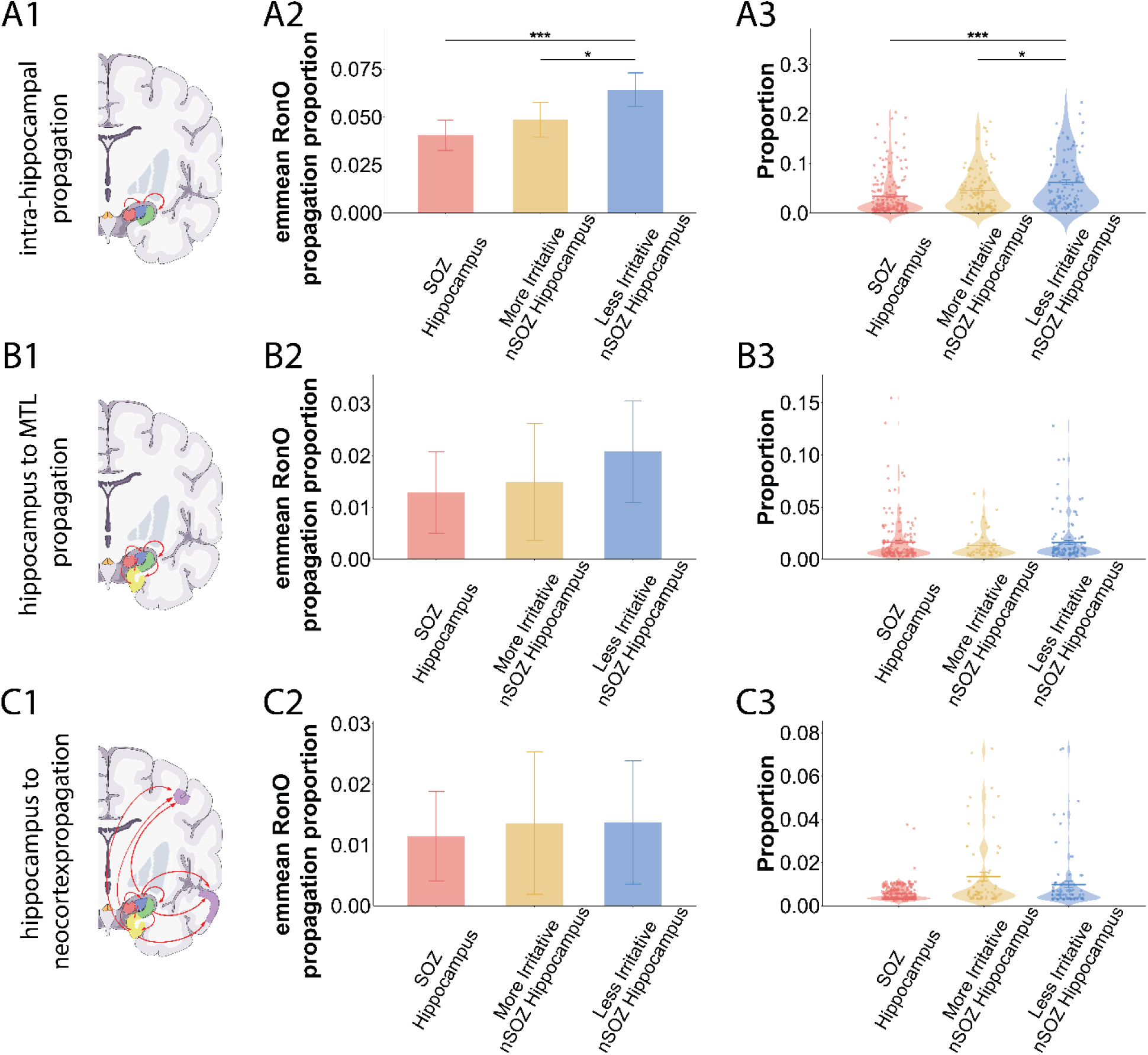
Analysis of RonO propagation proportion (i.e. probability) from the hippocampus, stratified by epileptogenicity, via two-way mixed-effects ANOVA. (A1-C1) schematic illustrations of hippocampal different propagation paths. (A1) Intra-hippocampal propagation, (B1) hippocampus to MTL propagation and (C1) and hippocampus to neocortex propagation. (A2-C2) bar plots showing the model-derived estimated marginal mean (emmean) of RonO propagation proportion and 95% CI across the three hippocampal groups using mixed-effects ANOVA. (A3-C3) Violin plots showing the actual RonO propagation proportions across three hippocampal groups with raw mean ± standard error. Each dot represents one observation. Post hoc pairwise comparisons were performed using the Holm–Bonferroni correction. Horizontal lines indicate significant pairwise differences between groups (*P<0.05, **P<0.01).

Next, we analyzed the propagation proportion of FRonOs. When constructing the temporal networks, pathypy and bootstrapping identified far fewer statistically significant propagation paths for FRonOs than for RonOs, precluding a direct and reliable comparison between the two. Furthermore, when evaluating the FRonO temporal networks using a two-way mixed-effects ANOVA, we found no significant interaction effect between group and category at maximum latencies of either 150 ms (Supplementary Table S5, p = 0.28) or 32.5 ms (Supplementary Table S6, p = 0.65). However, these results require cautious interpretation. The scarcity of FRonOs in Group 2 and Group 3 hippocampi (Fig. 2D2) makes this analysis inherently unbalanced and underpowered compared to our evaluation of RonOs.

## Discussion

In summary, in patients undergoing pre-surgical evaluation with SEEG, we stratified hippocampi into three groups based ictal and interictal epileptogenicity and constructed temporal networks to compare the statistically significant propagation proportions of RonO and fRonO in the iEEG across three propagation pathway categories: 1) intrahippocampal, 2) hippocampal to mesial-temporal, and 3) hippocampal to neocortical. Our primary statistically significant finding, with respect to HFO propagation, was that intrahippocampal RonO propagation proportion was highest in the least irritable non-SOZ hippocampi (Figure 5).

**Figure 5.**
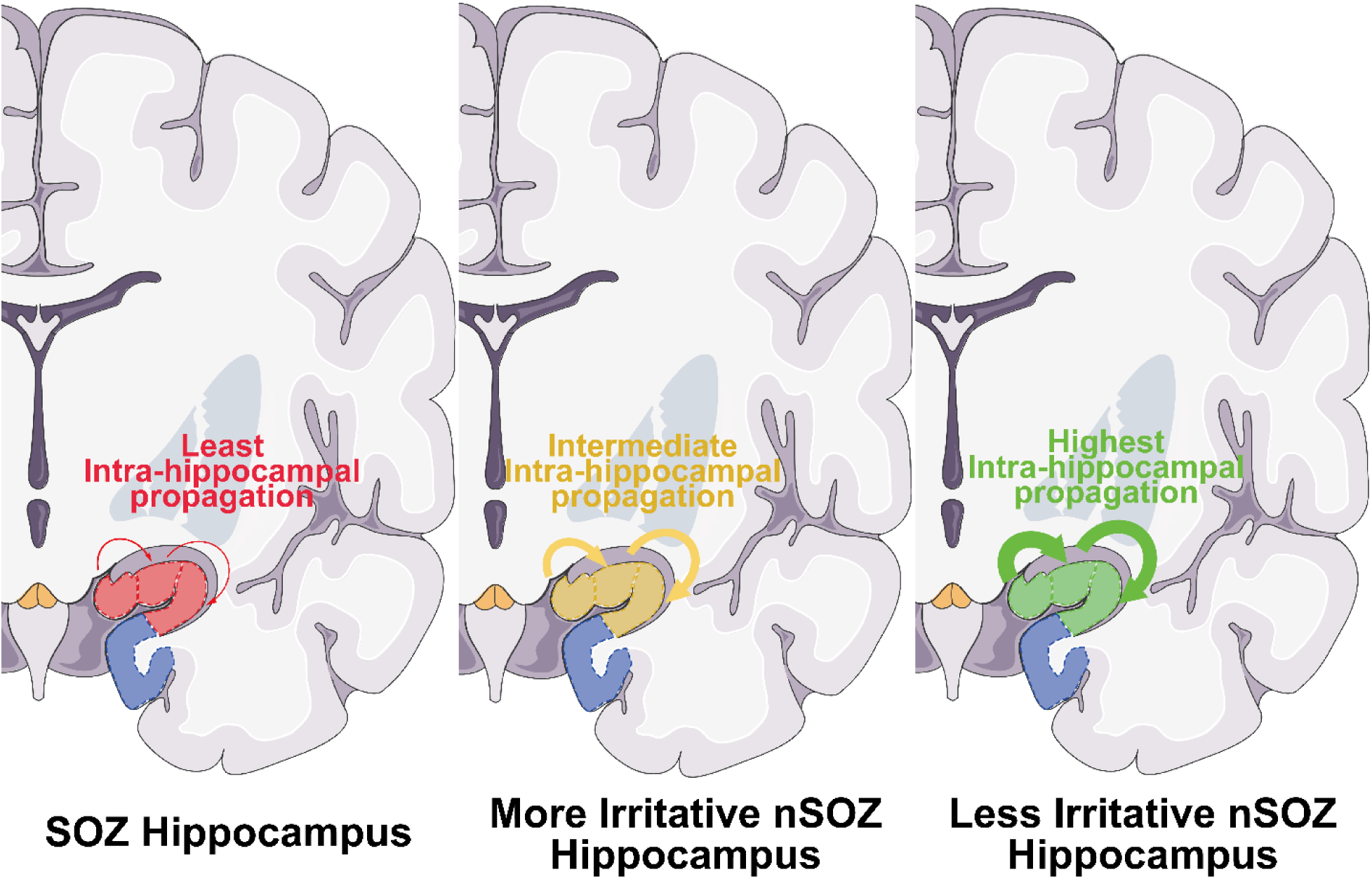
Schematic representation of intra-hippocampal ripple on oscillation (RonO) propagation along the spectrum of hippocampal epileptogenecity showing increased intrahippocampl RonO propagation in the less irritative hippocampus. The left panel represents the SOZ hippocampus, the middle panel represents the more irritative non-SOZ hippocampus, and the right panel represents the less irritative non-SOZ hippocampus. Colored overlays indicate the hippocampal region corresponding to each group, and curved arrows depict intra-hippocampal RonO propagation. Red, yellow, and green denote SOZ, more irritative non-SOZ, and less irritative non-SOZ hippocampus, respectively. The arrow thickness represent the porportion of propagation.

In this study, we constructed temporal networks with a maximum latency of <150 msec followed by bootstrapping to identify the statistically significant paths. This methodology is most suitable to identify the poly-synaptic pathways, or slower recruitment, associated with HFO propagation. This approach is less suitable for characterizing monosynaptic HFO propagation or HFO propagation associated with rapidly traveling wavefronts. Using a maximum latency of <32.5 msec to construct the temporal network we failed to observe statistically significant findings indicating that the significant increase in intrahippocampal RonO propagation proportion in the group of least irritative hippocampi could be attributed to the longer latency intrahippocampal polysynaptic RonO propagation paths. In patients undergoing presurgical evaluation with depth electrodes, intra-hippocampal ripple propagation with a latency up to 100 msec has been previously reported^42,43^.

In contrast to our observation that intrahippocampal propagation proportion is greatest in less irritative non-SOZ hippocampus, other studies have found that identifying the source node (i.e., iEEG recording contact), wherein the HFO generator originates and exhibits highest rate of HFO propagation, localizes the patient’s SOZ^33–36^. However, the methodologies of such studies differ from ours by considering HFO as a point process implementing automated event clustering, strict latency windows (up to <100 ms), and directed acyclic graphs to identify propagation clusters with a leader and a follower designed to map outward radiation from a single focal source. However, another study investigating HFO propagation in SEEG recordings used the sign test and reported that increased HFO rate was superior to increased HFO propagation in identifying the SOZ^44^.This conclusion is in accord with our observations that SOZ hippocampus did not exhibit any significant increase in RonO or FRonO propagation proportion.

Intrahippocampal ripple propagation proportion is highest in less-irritable hippocampus likely due to the preservation of micro-architecture, functional lateral inhibition, and intact homeostatic gating mechanisms^22^. Seizures originating in the hippocampus most often exhibit hypersynchronous (HYP) or low-voltage fast (LVF) onset patterns^45^. The HYP onset pattern remains mostly constrained to the hippocampus. Structural MRI investigations indicate that both onset types are associated with hippocampal injury and pathway disconnection; however, this damage is notably more pronounced in patients experiencing predominantly HYP onsets^46^. Presumably, this hippocampal injury and network disconnection drive the significant reduction in intrahippocampal RonO propagation observed in both the SOZ and the more irritative non-SOZ tissue.

We hypothesized that the proportion of physiological to pathological RonO exists on a spectrum tied to the severity of hippocampal injury. To test this, we examined whether ripple propagation properties across three categories of irritative and epileptogenic hippocampi could differentiate these underlying mixtures. Discriminating individual physiological and pathological RonO events is a known challenge. Prior comparisons of RonO spectral content, power, duration, and phase-amplitude coupling between SOZ and non-SOZ contacts yield statistically significant differences yet exhibit small effect sizes and high overlap^2,23,26^. While one interpretation of our findings is that intrahippocampal propagation proportions can distinguish tissue with a higher proportion of physiological ripples, we propose an alternative framework: the ripple signals and the hippocampal substrates generating them are fundamentally intertwined. Rather than representing two discrete populations, all ripple events may lie on a single physiological-to-pathological continuum dictated by the degree of local structural and functional hippocampal injury.

### Limitations

A technical limitation of this study involves the Hilbert detector used for HFO detection, which defined the onset as the time when the HFO reached half-maximal amplitude. While this inherent delay in onset time was likely consistent across most events, HFOs with non-uniform amplitude contours could introduce errors in the measurement of HFO onset time. Future work should validate these findings using HFO detectors utilizing wavelet transforms that more accurately define the time of HFO onset^47^. Other possible confounds include analyzing just a single segment of non-REM sleep^48^ and not examining patient outcomes with respect to resection of the hippocampal SOZ^35^. Although FRonO rates were significantly higher in the hippocampal SOZ, undetected epileptogenicity in the contralateral hippocampus may have compromised our analysis of differences in RonO and FRonO.

### Conclusions

In this study we characterized RonO and FRonO onset times measured in the iEEG from patients undergoing presurgical evaluation and utilized temporal networks validated with bootstrapping to identify statistically significant propagation paths of HFOs originating in the hippocampus and projecting in the ipsilateral hemisphere. We found that intrahippocampal RonO propagation proportion was significantly higher for least irritative hippocampi in the non-SOZ and were relatively longer latency and likely polysynaptic. We failed to demonstrate increased HFO propagation proportion from contacts in the hippocampal SOZ. Our findings suggest that either physiological hippocampal RonO have a higher intrahippocampal propagation proportion, or that individual hippocampal RonO are neither distinctly physiological or pathological but reflect the structural and functional integrity of their hippocampal substrate generator. Future investigations of RonO propagation in the epileptogenic hippocampus could utilize the neuropixel high density electrode array^49^ to better characterize possible monosynaptic RonO propagation and rapidly propagating wavefronts^28,50^.

## Data Availability

All data produced in the present study are available upon reasonable request to the authors

## Acknowledgments

This work was supported by the National Natural Science Foundation of China (grant numbers: 82171437, 82471469, and 82301636) and the Natural Science Foundation of Zhejiang Province (grant numbers: LD24H090003 and LY24H090004) his work was fully supported by the National Institute of Health K23 NS094633, a Junior investigator Award from the American Epilepsy Society (S.A.W.), R01 NS106957(R.J.S.), Resnick family foundation (R.S.), and the Christina Louise George Trust (R.J.S..,

## Ethical Approval

The study followed the Declaration of Helsinki and local ethical guidelines.

## Data Availability Statement

The data supporting the findings can be accessed upon reasonable request.

CRediT: Y.C (conceptualization, methodology, software, validation, formal analysis, investigation, data curation, writing – original draft and review and editing, visualization),H.Y (software, resources, data curation), L.H (software, resources, data curation), C.C (software, resources, data curation), R.S. (writing –review and editing), S.W. (conceptualization, investigation, writing –review and editing, supervision, project administration, funding acquisition), S.A.W (conceptualization, methodology, nvestigation, data curation, writing – original draft and review and editing, supervision, funding acquisition)

