## Supplementary Material for "Epileptogenicity alters intrahippocampal ripple propagation"

### Supplementary Methods

#### HFO detection

##### Preprocessing

The iEEG signals were preprocessed by filtering, normalization, and outlier data removed to reduce artifacts and enhance HFO events relative to the background. Raw signals were FIR bandpass-filtered in the ripple range of 80–250 Hz or fast-ripple range of 250–500 Hz. 0.5% of outlier data was removed from the normalized filtered signals to facilitate subsequent analysis.

##### Feature Extraction

We mainly focused on time and frequency domain of the iEEG signals. In a signal of length N: $X = (x_1, x_2, \ldots, x_N)$, where six features were extracted: Short-time Energy, Short-time Line Length, Bandpass Envelope, Hilbert Transform, Teager Energy Operator and Coastline Index-based Signal to Noise Ratio (CI-SNR).

1. Short-time Energy

For sample $x_t$ at time $t$ ($1 < t < \mathrm{N} + 1 - k$) in iEEG signals, the short-time energy $E_t$ can be calculated as:

$$E_t = \sqrt{\frac{1}{k}\sum_{i=t}^{t+k-1} x_i^2}$$

where $k$ is the length of the sliding window.

2. Short-time Line Length

Because short-time line length is more effective than short-time energy in reducing false-positive detections caused by high-amplitude spikes and artifacts, the short-time line length $L_t$at time $t(1 < t < N + 2 - k)$was used as the time-domain feature for HFO event detection:

$$E_t = \sqrt{\frac{1}{k}\sum_{i=t}^{t+k-2} |x_i - x_{i-1}|}$$

where $k$ is the length of the sliding window.

3. Bandpass Envelope

To characterize the amplitude profile of each HFO event, the local maxima of the band-pass filtered signal $x_H$ were extracted as the band-pass envelope feature:

$$ENV_t = \max\left(x_{H\left(t-\frac{k}{2}\right)}, \dots, x_{H\left(t+\frac{k}{2}\right)}\right)$$

4. Hilbert Transform

The Hilbert transform was used to quantify detect the energy increase of high frequency signals during HFO events and was defined as follows:

$$H[x_t] = \frac{1}{\pi} \int \frac{x_\tau}{t - \tau} dt$$

5. Teager Energy Operator

A non-linear energy operator that reflects the instantaneous frequency and amplitude information of iEEG signals and was defined as follows:

$$\phi(x_t) = x_t x_{t+k-2} - x_{t-1} x_{t+k-1}$$

When k=2, the upper formula is the Teager energy operator.

6. Coastline Index-based Signal to Noise Ratio (CI-SNR)

To reduce the influence of low-frequency activity and emphasize HFO-related features, the high-frequency band containing HFOs was defined as the target frequency band, whereas the low-frequency component between 0 and 80 Hz was defined as the noise frequency band. The frequency dominance signal to noise ratio (SNR) was calculated as follows:

$$FD_{SNR_t} = \sqrt{\sum_{i=t-k/2}^{t+k/2} x_{H(i)}^2 \Big/ \sum_{i=t-k/2}^{t+k/2} x_{L(i)}^2}$$

where $k$ is the length of the sliding window; $x_H$ is the target frequency band signal after FIR filtering; and $x_L$ is the noise frequency band signal after FIR filtering.

To avoid the disturbance of low frequency spikes and represent the multi-spike characteristics of HFOs in a better way, we proposed a CI-SNR:

$$CI - SNR_t = 20lg\left(\sqrt{\sum_{i=t-k/2}^{t+k/2}\left(x_{H(i)} - x_{H(i-1)}\right)^2 / \sum_{i=t-k/2}^{t+k/2}\left(x_{L(i)} - x_{L(i-1)}\right)^2}\right)$$

Feature values were averaged within a 10-ms sliding window. For short-time energy, short-time line length, the Teager energy operator, and the Hilbert transform feature, detection thresholds were defined as the upper 25% of feature values calculated across all channels for each subject. For the CI-SNR and band-pass envelope features, thresholds were defined as the mean feature value across all channels for each subject. Samples with feature values exceeding the corresponding threshold were considered candidate HFO events for that feature.

$$HFO_{feature} = \frac{(sign(feature - feature_{threshold}) + 1)}{2}$$

$$Multi\_domain\ features\ based\ HFO = sign(feature_{TEO} + feature_{bandpass\ envelop} +$$

$$0.25feature_{short\_time\ energy} + 0.25feature_{short\_time\ line\ length} +$$

$$0.25feature_{hilbert\ transform} + 0.25feature_{CI\_SNR} - 1).$$

**Artifact Elimination**

Oscillation events were rejected as artifacts if their peak-to-valley amplitude was less than three times the average peak-to-valley amplitude of the neighboring environmental range, or if more than 10 consecutive oscillations were present in the neighboring environmental range. In addition, events were excluded if their duration was outside the 10–500 msec range, or oscillation amplitude less than the average value plus 4SD, or center frequency < 80 Hz were also excluded.

### IED detection and RonO / FRonO identification

**HFO detection**

Interictal epileptiform discharges (IEDs) were automatically detected using the signal-envelope distribution modeling method described by Janca et al. Signals acquired at higher sampling rates were first resampled to 200 Hz to maintain consistent filter characteristics. Each channel was then zero-phase band-pass filtered between 10 and 60 Hz using a combination of eighth-order Type II Chebyshev high-pass and low-pass filters. Power-line interference at 50Hz was attenuated using a biquadratic notch filter with poles positioned on a circle of radius 0.985 and a 4-Hz stopband.

For each filtered signal, the instantaneous signal envelope was calculated from the absolute value of its Hilbert transform. Then the envelope was analyzed using 5s moving windows with 80% overlap between consecutive windows. Within each window, the distribution of the envelope amplitudes was modeled as a log-normal distribution using maximum-likelihood estimation. Its probability density function was:

$$y = f(x|\mu, \sigma) = \frac{1}{x\sigma\sqrt{2\pi}} e^{-\frac{(\ln x - \mu)^2}{2\sigma^2}}; x > 0$$

where $y$ is probability density, and $x$ represents the envelope amplitude, and $\mu$ and $\sigma$ are the mean and standard deviation of the logarithmically transformed envelope values. For an envelope segment containing (N) samples, these parameters were estimated as:

$$\mu = \frac{1}{N}\sum_{i=1}^{N} \ln(x_i)\,, \sigma = \sqrt{\frac{1}{N-1}\sum_{I=1}^{N}[\ln(x_i - \mu)]^2}$$

IED-containing segments are characterized by a positively skewed envelope distribution with increased variance and a long right tail. Therefore, an adaptive detection threshold was calculated from the mode and median of the fitted log-normal distribution:

$$th = k_1[Mode + Median];\ Mode = e^{\mu-\sigma^2}, Median = e^{\mu}$$

where $\mu$ and $\sigma$ were estimated using MLE.

**RonO / FRonO identification**

The IED detection toolbox identified the peak time of each IED. An HFO was classified as a RonO or FRonO if no IED peak occurred between its onset and offset.

### HFO temporal network construction and propagation analysis

Time-respecting path is a sequence of time-stamped edges that respect the time order of interactions in a temporal network[39] . Given a sequence of node and their occurring time: $(v_0, t_0)$, $(v_1, t_1)$, $(v_2, t_2)$, …, $(v_{n-1}, t_{n-1})$, $(v_n, t_n)$ $(t_0 < t_1 < t_2 < \cdots < t_{n-1} < t_n)$, the interaction $(v_i, v_j; t_i)$ $(i < j,\ i, j \in (0,1,2, \ldots, n-1))$ exists if the time difference:

$$t_j - t_i < \varepsilon,$$

where $\varepsilon$ denotes the maximum latency. Using this rule, we defined a time-respecting path from source node $v_0$ to target node $v_n$ with the following interactions:

$$(v_0, v_1; t_0), (v_1, v_2; t_1), (v_2, v_3; t_2), \ldots, (v_{n-1}, v_n; t_{n-1})$$

where $t_0 < t_1 < t_2 < \cdots < t_{n-1}$. In order to strengthen the real connection between source node and target node, we limited the maximum time gap between adjacent time-stamped links $(v_{i-1}, v_i; t_{i-1})$, $(v_i, v_{i+1}; t_i)$ to $\delta$ that $0 < t_i - t_{i-1} < \delta$ $(i \in (1,2, \ldots, n-1))$. In this case, $\delta$ was equal to maximum latency. This step was calculated using python package *pathpy* (https://github.com/pathpy/pathpy). HFO propagation was identified for ripple on oscillation/background (i.e., RonO 80-250 Hz) and fast RonO (FRonO i.,e, 250-600 Hz) separately. Pathpy was used to generate two distinct temporal networks for both RonO and fRonO using a maximum latency ($\varepsilon$) of 150 msec and 32.5 msec.

To control for chance HFO propagation, we applied a permutation test to the distinct RonO and fRonO temporal networks. For each hemisphere, we generated 1,000 surrogate networks by randomizing HFO onset times within each node (depth electrode contact) while preserving total event counts. After processing each surrogate through pathpy to count HFO sequences along time-respecting paths, we standardized the actual (i.e., empirical) path counts by calculating z-scores relative to the mean and standard deviation of the surrogate distributions. This process was repeated using a maximum latency (ε) of 150 msec and 32.5 msec

$$z - score = \frac{n_{actual} - \mu_{surrogate}}{\sigma_{surrogate}}$$

A propagation with:

$$z - score > z_{min}$$

were retained, which mean we should estimate the minimum z using combinatorics. The median number of contacts included in the analysis was 96, and hops up to 5 could capture most of the propagations. According to this information, first we calculated the combinatorial space. In a temporal graph network with n = 96 nodes, the number of possible paths were calculated as follows:

number of 1hop path (2 nodes): $n \times (n-1) = 9120$;

number of 2hops path (3 nodes): $n \times (n-1)^2 = 866400$;

number of 3hops path (4 nodes): $n \times (n-1)^3 = 8230800$;

number of 4hops path (5 nodes): $n \times (n-1)^4 = 7819260000$;

number of 5hops path (6 nodes): $n \times (n-1)^5 = 742829700000$;

The total combinatorial search space: $\mathrm{M} = 750732143520$.

Then we calculated the adjusted false-positive rate ($\alpha$= 0.05) using Bonferroni correction:

$$\alpha_{adj} = \frac{0.05}{750732143520} \approx 6.66 \times 10^{-14}$$

Finally, converted this p-value into a z-score: $z_{min} \approx 7.40$. As a result, it was reasonable to set $z_{min}$ as 7.5 in this research. After the cut-off z-score, there were still some propagations with a z-score > 7.5 and a very low frequency of occurrence, which in most cases the distribution of these propagations in the surrogated data were zero distribution. These propagations with low frequency were also considered as random occurred. As a result, propagations with a frequency > 3 times/hour were retained.

Propagation proportion was defined as, for a given hippocampal channel, the probability that an HFO event recorded on that hippocampal channel participates in a propagation path (to another channel).

$$Proportion = \frac{count\ of\ a\ path\ from\ a\ given\ hippocampal\ channel}{count\ of\ HFO\ events\ in\ this\ hippocampal\ channel}$$

Last, to reduce dimensionality, remaining propagations were categorized into 3 categories based on the propagating area: 1) intra-hippocampal propagation; 2) hippocampus to MTL propagation; 3) hippocampus to neocortex propagation. Importantly, Importantly, if a path originating from the hippocampus made a hop to the mesial-temporal lobe (MTL) or neocortex and then a subsequent hop back to the hippocampus, by convention the path was categorized as hippocampus to MTL, or hippocampus to neocortex, respectively.

### Supplementary Data

standard error of the mean. Numbers inside the bars indicate the group mean.

**Aligned Rank Transform mixed-effects ANOVA**

| **Term** | **F** | **Df** | **Df.res** | **Pr(>F)** |
|---|---|---|---|---|
| Group | 0.059712 | 2 | 63.653 | 0.94209 |

**Aligned-rank estimated marginal mean**

| **group** | **emmean** | **SE** | **df** | **lower.CL** | **upper.CL** |
|---|---|---|---|---|---|
| **Less Irritative nSOZ Hippocampus** | 101.9868 | 11.8534 | 62.89 | 78.2989 | 125.6747 |
| **More Irritative nSOZ Hippocampus** | 107.0114 | 10.7075 | 62.37 | 85.6100 | 128.4128 |
| **SOZ Hippocampus** | 102.9613 | 9.6831 | 66.12 | 83.6291 | 122.2935 |

Table S1. Aligned Rank Transform mixed-effects ANOVA for RonO rates (events/min) across groups. Hippocampal RonO rates (events/min) were compared among 3 groups (SOZ Hippocampus, More Irritative nSOZ Hippocampus and Less Irritative nSOZ Hippocampus) using Aligned Rank Transform mixed-effects ANOVA.

**Aligned Rank Transform mixed-effects ANOVA**

| Term | F | Df | Df.res | Pr(>F) |
|---|---|---|---|---|
| Group | 28.259 | 2 | 62.101 | 1.875e-09 |

**Aligned-rank estimated marginal mean**

| group | emmean | SE | df | lower.CL | upper.CL |
|---|---|---|---|---|---|
| **Less Irritative nSOZ Hippocampus** | 51.815580 | 8.753005 | 61.85 | 34.31774 | 69.31343 |
| **More Irritative nSOZ Hippocampus** | 101.781330 | 7.890218 | 60.14 | 85.99932 | 117.56335 |
| **SOZ Hippocampus** | 137.054380 | 7.207935 | 64.71 | 122.65790 | 151.45086 |

**Post hoc comparison**

| Comparison | Estimate | SE | df | t.ratio | p.value |
|---|---|---|---|---|---|
| **Less Irritative** vs **More Irritative** | -49.96575 | 11.78434 | 61.08 | -4.24 | 0.0002 |
| **Less Irritative** vs **SOZ** | -85.23879 | 11.33884 | 62.98 | -7.517 | <.0001 |
| **More Irritative** vs **SOZ** | -35.27304 | 10.6869 | 62.16 | -3.301 | 0.0016 |

Table S2. Aligned Rank Transform mixed-effects ANOVA for FRonO rates (events/min) across groups. Hippocampal FRonO rates (events/min) were compared among 3 groups (SOZ Hippocampus, More Irritative nSOZ Hippocampus and Less Irritative nSOZ Hippocampus) using Aligned Rank Transform mixed-effects ANOVA. Post hoc pairwise comparisons were performed using **aligned-rank estimated marginal means** with Holm–Bonferroni correction for multiple comparisons.

**Mixed-effects ANOVA**

**Response: RonO propagation (maximum latency 150msec) proportion**

| **Term** | **Chisq** | **Df** | **Pr(>Chisq)** |
|---|---|---|---|
| (Intercept) | 142.5636 | 1 | < 2.2e-16 |
| group | 4.6713 | 2 | 0.096748 |
| category | 247.5308 | 2 | < 2.2e-16 |
| group:category | 14.4663 | 4 | 0.005946 |

**Estimated marginal mean**

| **group** | **category** | **emmean** | **SE** | **df** | **lower.CL** | **upper.CL** |
|---|---|---|---|---|---|---|
| **Less Irritative nSOZ Hippo** | **Intra-hippocampal propagation** | 0.06420 | 0.00441 | 81.9 | 0.05545 | 0.07300 |
| **More Irritative nSOZ Hippo** | **Intra-hippocampal propagation** | 0.04850 | 0.00457 | 74.9 | 0.03940 | 0.05760 |
| **SOZ Hippo** | **Intra-hippocampal propagation** | 0.04050 | 0.00398 | 102.1 | 0.03260 | 0.04840 |
| **Less Irritative nSOZ Hippo** | **Hippocampus to MTL propagation** | 0.02080 | 0.00492 | 95.5 | 0.01100 | 0.03050 |
| **More Irritative nSOZ Hippo** | **Hippocampus to MTL propagation** | 0.01490 | 0.00569 | 168.9 | 0.00367 | 0.02620 |
| **SOZ Hippo** | **Hippocampus to MTL propagation** | 0.01290 | 0.00395 | 95.2 | 0.00507 | 0.02080 |
| **Less Irritative nSOZ Hippo** | **Hippocampus to neocortex propagation** | 0.01380 | 0.00513 | 120.6 | 0.00359 | 0.02390 |
| **More Irritative nSOZ Hippo** | **Hippocampus to neocortex propagation** | 0.01360 | 0.00594 | 138.6 | 0.00188 | 0.02540 |
| **SOZ Hippo** | **Hippocampus to neocortex propagation** | 0.01150 | 0.00372 | 75.8 | 0.00406 | 0.01890 |

**Post hoc comparison**

| **Category** | **Comparison** | **Estimate** | **SE** | **df** | **t.ratio** | **p.value** |
|---|---|---|---|---|---|---|
| **Intra-hippocampal propagation** | **Less Irritative nSOZ –More Irritative nSOZ** | 0.015722 | 0.006350 | 78.2 | 2.477 | 0.0308 |
| **Intra-hippocampal propagation** | **Less Irritative nSOZ – SOZ** | 0.023725 | 0.005940 | 90.2 | 3.996 | 0.0004 |

| | | | | | | |
|---|---|---|---|---|---|---|
| **Intra-hippocampal propagation** | **More Irritative nSOZ – SOZ** | 0.008002 | 0.006060 | 85.2 | 1.321 | 0.19 |
| **Hippocampus to MTL propagation** | **Less Irritative nSOZ –More Irritative nSOZ** | 0.005853 | 0.007530 | 130.1 | 0.778 | 0.8765 |
| **Hippocampus to MTL propagation** | **Less Irritative nSOZ – SOZ** | 0.007848 | 0.006310 | 95.4 | 1.243 | 0.6505 |
| **Hippocampus to MTL propagation** | **More Irritative nSOZ – SOZ** | 0.001996 | 0.006930 | 138.0 | 0.288 | 0.8765 |
| **Hippocampus to neocortex propagation** | **Less Irritative nSOZ –More Irritative nSOZ** | 0.000139 | 0.007850 | 130.4 | 0.018 | 1 |
| **Hippocampus to neocortex propagation** | **Less Irritative nSOZ – SOZ** | 0.002283 | 0.006340 | 101.6 | 0.360 | 1 |
| **Hippocampus to neocortex propagation** | **More Irritative nSOZ – SOZ** | 0.002144 | 0.007010 | 114.9 | 0.306 | 1 |

Table S3. Two-way mixed-effect ANOVA for RonO propagation proportions and post hoc pairwise comparisons. Maximum latency was set as 150msec to generate RonO propagation from hippocampus. Then a mixed-effect ANOVA was used to test the effect of group (SOZ Hippocampus, More Irritative nSOZ Hippocampus and Less Irritative nSOZ Hippocampus) and category (intra-hippocampal propagation, hippocampus to MTL propagation, and hippocampus to neocortical propagation) and their interaction (group × category) effect on RonO propagation proportion. Post hoc comparison was performed across 4 groups within each category based on estimated marginal means using Holm-Bonferroni multiple comparison correction.

**Mixed-effects ANOVA**

**Response: RonO propagation (maximum latency 32.5msec) proportion**

| Term | Chisq | Df | Pr(>Chisq) |
|---|---|---|---|
| (Intercept) | 116.5415 | 1 | <2e-16 |
| Group | 2.2148 | 2 | 0.3304 |
| category | 255.9929 | 2 | <2e-16 |
| group:category | 6.2022 | 4 | 0.1845 |

**Estimated marginal mean**

| group | Category | emmean | SE | df | lower.CL | upper.CL |
|---|---|---|---|---|---|---|
| **Less Irritative nSOZ Hippo** | **Intra-hippocampal propagation** | 0.07220 | 0.00598 | 70.2 | 0.06023 | 0.08410 |
| **More Irritative nSOZ Hippo** | **Intra-hippocampal propagation** | 0.06520 | 0.00594 | 71.1 | 0.05333 | 0.07700 |
| **SOZ Hippo** | **Intra-hippocampal propagation** | 0.05380 | 0.00498 | 74.2 | 0.04386 | 0.06370 |
| **Less Irritative nSOZ Hippo** | **Hippocampus to MTL propagation** | 0.02830 | 0.00665 | 87.4 | 0.01504 | 0.04150 |
| **More Irritative nSOZ Hippo** | **Hippocampus to MTL propagation** | 0.01860 | 0.00677 | 111.2 | 0.00516 | 0.03200 |
| **SOZ Hippo** | **Hippocampus to MTL propagation** | 0.02000 | 0.00536 | 90.5 | 0.00936 | 0.03070 |
| **Less Irritative nSOZ Hippo** | **Hippocampus to neocortex propagation** | 0.02390 | 0.00796 | 181.4 | 0.00823 | 0.03960 |
| **More Irritative nSOZ Hippo** | **Hippocampus to neocortex propagation** | 0.01160 | 0.00835 | 167.6 | -0.00488 | 0.02810 |
| **SOZ Hippo** | **Hippocampus to neocortex propagation** | 0.01780 | 0.00699 | 226.6 | 0.00405 | 0.03160 |

Table S4. Two-way mixed-effect ANOVA for RonO propagation proportions. Maximum latency was set as 32msec to generate RonO propagation from hippocampus. Then a mixed-effect ANOVA was used to test the effect of group (SOZ Hippocampus, More Irritative nSOZ Hippocampus and Less Irritative nSOZ Hippocampus) and category (intra-hippocampal propagation, hippocampus to MTL propagation, and hippocampus to neocortical propagation) and their interaction (group × category) effect on RonO propagation proportion.

| **Mixed-effects ANOVA** | | | |
|---|---|---|---|
| **Response: FRonO propagation (maximum latency 150msec) proportion** | | | |
| **Term** | **Chisq** | **Df** | **Pr(>Chisq)** |
| (Intercept) | 23.8576 | 1 | 0.00000 |
| group | 5.8619 | 2 | 0.05335 |
| category | 7.2650 | 2 | 0.02645 |
| group:category | 1.6550 | 3 | 0.64697 |

| **Estimated marginal mean** | | | | | | |
|---|---|---|---|---|---|---|
| **group** | **category** | **emmean** | **SE** | **df** | **lower.CL** | **upper.CL** |
| **Less Irritative nSOZ Hippo** | **Intra-hippocampal propagation** | 0.1550 | 0.0367 | 47.0 | 0.0811 | 0.2289 |
| **More Irritative nSOZ Hippo** | **Intra-hippocampal propagation** | 0.1408 | 0.0206 | 46.6 | 0.0994 | 0.1822 |
| **SOZ Hippo** | **Intra-hippocampal propagation** | 0.0720 | 0.0167 | 39.5 | 0.0382 | 0.1058 |
| **Less Irritative nSOZ Hippo** | **Hippocampus to MTL propagation** | 0.0986 | 0.0516 | 141.5 | -0.0035 | 0.2006 |
| **More Irritative nSOZ Hippo** | **Hippocampus to MTL propagation** | 0.0853 | 0.0312 | 157.6 | 0.0237 | 0.1469 |
| **SOZ Hippo** | **Hippocampus to MTL propagation** | 0.0430 | 0.0172 | 43.5 | 0.0084 | 0.0777 |
| **Less Irritative nSOZ Hippo** | **Hippocampus to neocortex propagation** | nonEst | NA | NA | NA | NA |
| **More Irritative nSOZ Hippo** | **Hippocampus to neocortex propagation** | 0.1415 | 0.0437 | 299.0 | 0.0554 | 0.2276 |
| **SOZ Hippo** | **Hippocampus to neocortex propagation** | 0.0451 | 0.0221 | 107.8 | 0.0013 | 0.0888 |

Table S5. Two-way mixed-effect ANOVA for FRonO propagation proportions. Maximum latency was set as 150msec to generate FRonO propagation from hippocampus. Then a mixed-effect ANOVA was used to test the effect of group (SOZ Hippocampus, More Irritative nSOZ Hippocampus and Less Irritative nSOZ Hippocampus) and category (intra-hippocampal propagation, hippocampus to MTL propagation, and hippocampus to neocortical propagation) and their interaction (group × category) on FRonO propagation proportion.

| Mixed-effects ANOVA | | | | | | |
|---|---|---|---|---|---|---|
| **Response: FRonO propagation (maximum latency 32msec) proportion** | | | | | | |
| **Term** | **Chisq** | | **Df** | | **Pr(>Chisq)** | |
| (Intercept) | 6.5494 | | 1 | | 0.0104917 | |
| Group | 0.0854 | | 2 | | 0.958212 | |
| category | 15.3605 | | 2 | | 0.0004619 | |
| group:category | 3.8470 | | 3 | | 0.278472 | |
| **Estimated marginal mean** | | | | | | |
| **group** | **category** | **emmean** | **SE** | **df** | **lower.CL** | **upper.CL** |
| **Less Irritative nSOZ Hippo** | **Intra-hippocampal propagation** | 0.15317 | 0.01880 | 62.80000 | 0.11550 | 0.19080 |
| **More Irritative nSOZ Hippo** | **Intra-hippocampal propagation** | 0.10886 | 0.00999 | 54.40000 | 0.08880 | 0.12890 |
| **SOZ Hippo** | **Intra-hippocampal propagation** | 0.07276 | 0.00766 | 39.20000 | 0.05730 | 0.08830 |
| **Less Irritative nSOZ Hippo** | **Hippocampus to MTL propagation** | 0.12030 | 0.03190 | 207.20000 | 0.05750 | 0.18310 |
| **More Irritative nSOZ Hippo** | **Hippocampus to MTL propagation** | 0.05376 | 0.01770 | 163.30000 | 0.01890 | 0.08860 |
| **SOZ Hippo** | **Hippocampus to MTL propagation** | 0.04260 | 0.00886 | 62.70000 | 0.02490 | 0.06030 |
| **Less Irritative nSOZ Hippo** | **Hippocampus to neocortex propagation** | nonEst | NA | NA | NA | NA |
| **More Irritative nSOZ Hippo** | **Hippocampus to neocortex propagation** | -0.00755 | 0.05850 | 384.50000 | -0.12260 | 0.10750 |
| **SOZ Hippo** | **Hippocampus to neocortex propagation** | 0.04759 | 0.01850 | 189.20000 | 0.01110 | 0.08410 |

Table S6. Two-way mixed-effect ANOVA for FRonO propagation proportions. Maximum latency was set as 32msec to generate FRonO propagation from hippocampus. Then a mixed-effect ANOVA was used to test the effect of group (SOZ Hippocampus, More Irritative nSOZ Hippocampus and Less Irritative nSOZ Hippocampus) and category (intra-hippocampal propagation, hippocampus to MTL propagation, and hippocampus to neocortical propagation) and their interaction (group × category) effect on FRonO propagation proportion.
